# Stool Genomics for Enhanced Monitoring of Nutrition Interventions in Childhood Malnutrition

**DOI:** 10.1101/2025.06.04.25328947

**Authors:** Ammara Aqeel, Najeeha T. Iqbal, Sanam Iram Soomro, Sheraz Ahmed, Teresa K. McDonald, Olivia Osborne, Sharon Jiang, Nolan Ives, Kazi Ahsan, Fayaz Umrani, Michael J. Barrat, Jeffrey I. Gordon, S. Asad Ali, Lawrence A. David

## Abstract

Ready-to-use therapeutic and supplementary foods (RUTF/RUSF) are a primary treatment for childhood malnutrition, but measuring intervention compliance is labor intensive. We applied FoodSeq, a fecal genomic dietary assessment biomarker, in malnourished infants (3-15 months) from Matiari, Pakistan. FoodSeq identified a significant spike in chickpea, a primary RUSF ingredient, during RUSF administration, highlighting the potential of genomics as an empirical tool for compliance monitoring and dietary analysis in community-based malnutrition programs.

## Main Text

Childhood malnutrition affected over 194 million children in 2022 and accounted for 45% of deaths in children under five globally^1^. Driven by a combination of socio-economic, environmental, and biologic factors^2,3,4,5,6^, malnutrition disproportionately impacts low- and middle-income countries (LMICs), leading to long-term health, social, and economic consequences^1,7^. Global treatment regimens utilize Community-Based Management of Acute Malnutrition (CMAM), deploying community health workers (CHWs) to screen and enroll children with malnutrition in an outpatient treatment regimen^8,9^. For severe acute malnutrition (SAM; weight-for-height z-score (WHZ) <-3 standard deviations (SD) from median WHO growth standards or mid-upper arm circumference (MUAC) <115 mm^10^), a weekly prescription of weight-adjusted ready-to-use therapeutic food (RUTF) and systematic antibiotic treatment is provided ^8,9,11^. For moderate acute malnutrition (MAM; WHZ between -2 and -3 SD or a MUAC between 115–125 mm^10^) ready-to-use supplemental food (RUSF) is provided and an antibiotic regimen is prescribed for underlying concomitant infection as needed^12^.

RUTF/SFs are formulated as a shelf-stable, single-dose foil package of an energy-nutrient dense paste eaten directly from the packet^13^. Although adherence to RUTF/SF therapy is crucial for adequate weight gain^14^, both socio-economic and dietary factors can impact treatment efficacy. For example, RUTF might be shared among household members or sold for supplementary income^15,16^, or age-inappropriate foods might be used for complementary feeding which can impair weight gain^17^. However, monitoring RUTF compliance involves regular CHW home-visits and/or collection of empty RUTF packets^8,9,14,18,19^, requiring extensive training and resources^20^. Novel approaches for objective monitoring of both RUTF/SF adherence and potential impacts of demographic, socioeconomic and/or dietary factors on compliance are needed. Dietary genomics, specifically FoodSeq, offer an objective, cost-effective alternative, characterizing dietary intake from residual food DNA in human stool^21^.

Here, we applied FoodSeq (*trnL-P6* chloroplast marker for plants^21^ and *12SV5* mitochondrial marker for vertebrates^22–24^) to a subset of an infant cohort from *Matiari*, Pakistan^18^. A total of 150 stool samples were included from 60 infants with MAM (3.1 – 15.2 months of age) and 30 healthy infants (12.1 – 15.2 months of age)^18^ (**Fig. 1a**). In this study, caregivers were provided breast-feeding and complementary feeding education at home for four weeks after enrollment (infants 3–6–months-of-age) (**Fig. 1a**). A locally produced RUSF *Acha Mum,* made from chickpea, edible oil, dried milk, and fortified with a vitamin and mineral mix^18,19^, was administered to wasted children of weaning age (mean age of 9.89 ± 0.99 months) for an average duration of 57.8 ± 6.3 days (**Fig. 1a**).

**Fig. 1.**
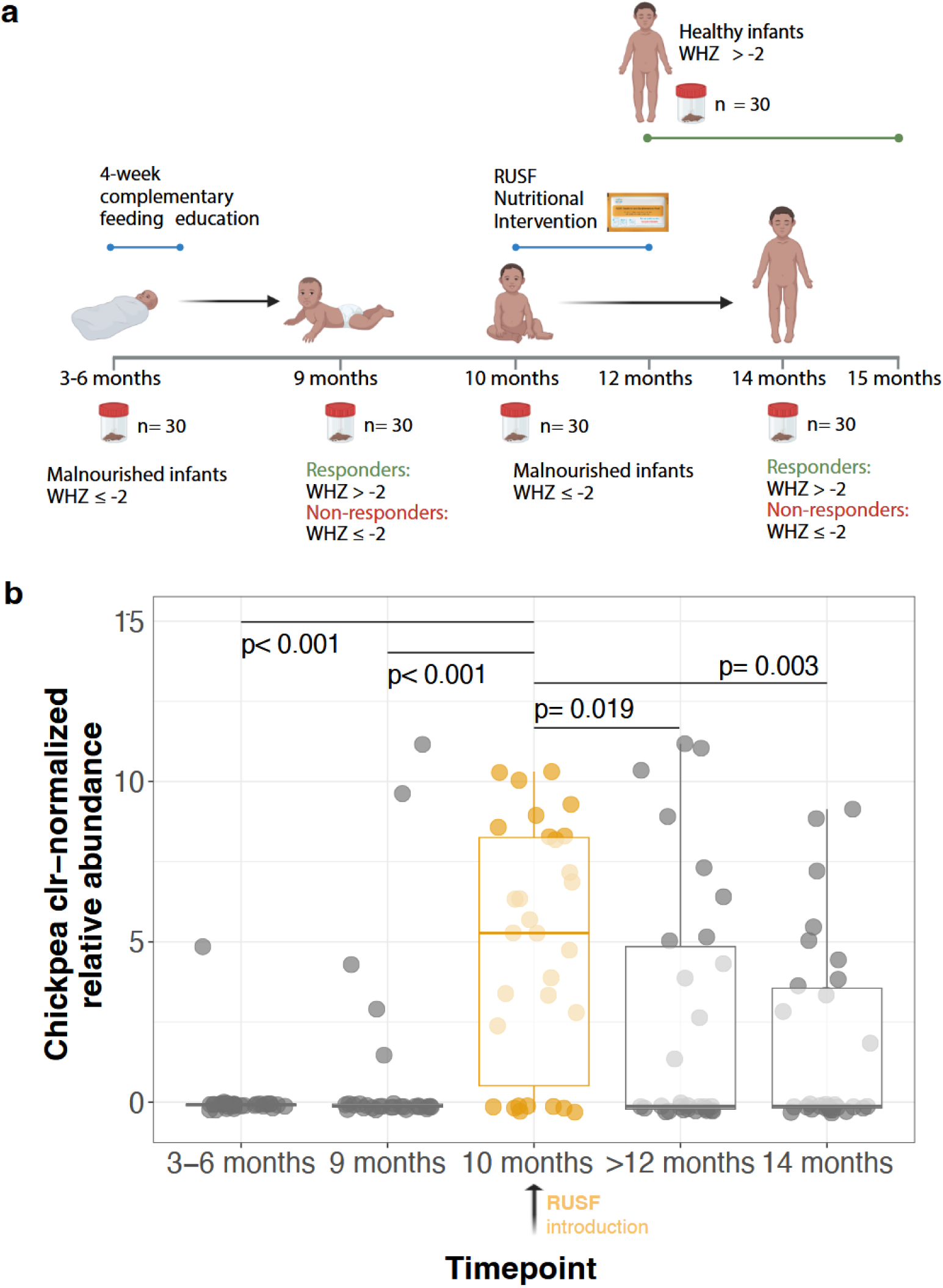
Sampling design of infant subset from SEEM cohort included in this study and detection of Chickpea DNA from stool samples aligning with administration of RUSF. **(a)** A cohort of 350 malnourished infants was followed longitudinally in the SEEM study through ages 3-6 months to 15 months ^18^. From this cohort, our study obtained stool samples from a subset of 30 infants with moderate acute malnutrition (MAM; weight-for-height z-scores (WHZ) ≤ -2) at 3–6 months of age when caregivers received an educational intervention covering WHO recommended breast feeding and complementary feeding practice and at approximately 9 months and another subset of 30 malnourished infants at approximately 10 months of age when caregivers were provided a locally-produced, chickpea-based RUSF (*Acha Mum*) for children and subsequently at approximately 14 months of age . Thirty stool samples from healthy infants (WHZ > 0, HAZ > −1.0) between the ages of 12.1–15.2 months were also included in the analysis. **(b)** CLR-normalized relative abundances of chickpea (*Cicer arietinum*) DNA amplified from stool samples compared across all timepoints. The 10-month timepoint when RUSF was administered is highlighted in orange. Box plots show median and interquartile ranges, while individual dots represent samples from each subject. *Wilcoxon signed-rank test* for 3-6 months vs 9 months and 10 months vs 14 months and *Wilcoxon rank-sum test* for all other timepoint comparisons were conducted. Significance results of statistical testing are marked.

FoodSeq detected a significant spike in chickpea DNA (*Cicer arietinum;* primary plant ingredient of *Acha Mum*) center-log-ratio normalized relative abundances (hereafter “CLR value”) coinciding with *Acha Mum* administration at 10 months in the MAM subset (median= 5.27, IQR [7.75]; present in 22/30 total samples, **Fig. 1b**). These values were significantly higher than those observed at pre-administration time points (median= -0.07, IQR [0.06] at 3-6 months, p < 0.001 and median= -0.13, IQR [0.10], p < 0.001 at 9 months; *Wilcoxon rank-sum test*), in healthy controls (median= -0.13, IQR [5.06], p = 0.019; *Wilcoxon rank-sum test*), and post-administration in MAM samples at 14 months (median= - 0.12, IQR [3.75], p = 0.003; *Wilcoxon signed-rank test*) (**Fig. 1b**). Other dietary plant taxa accounting for significant variation in dietary composition among participants (Principal Components Analysis (PCA): wheat; tea; banana; cowpea family; chickpea; carrot, parsley, cumin family; pea; wild rice; pepper family; coriander, cilantro, **Fig. 2a, Table S1**) did not display a similar spike during RUSF administration (**Fig. S1**).

**Fig. 2.**
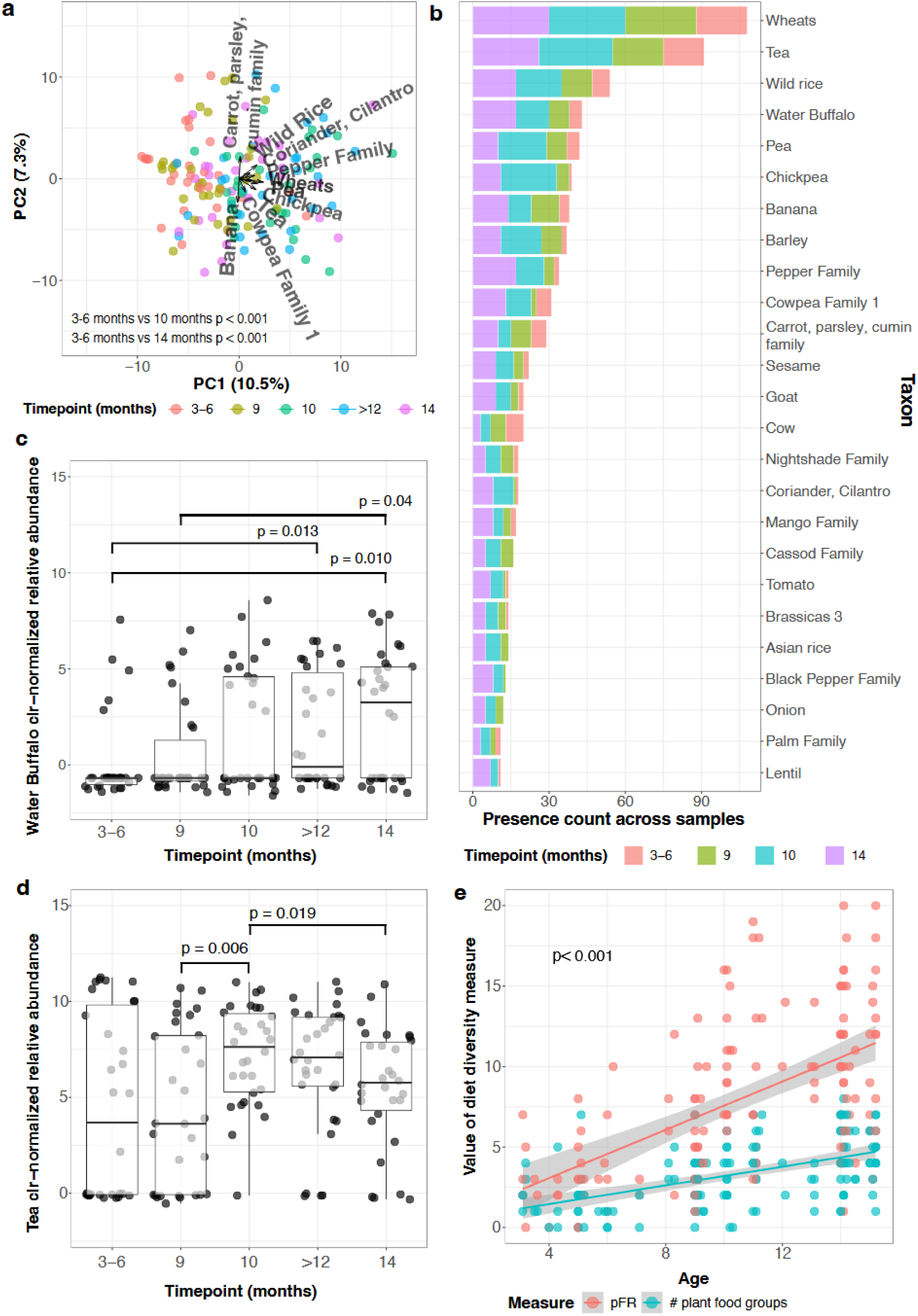
FoodSeq captures both regionally and developmentally expected dietary patterns as well as uncovers expected dietary behaviors relevant for intervention efficacy. (**a**) Principal Component Analysis (PCA) of plant dietary composition with biplot indicating relative contributions of top 10 dietary components. Points colored by timepoint (bottom of figure) and compared along PC1 values using linear mixed effect models with gender and PCR batch as fixed effects and sample ID as random effects. Significant differences as marked. (**b**) Bar plot showing the presence counts of top 25 food (plant and vertebrate combined) taxa detected in participant stool samples across all timepoints except >12 months (healthy controls, see Appendix B: Fig. 14b for top 25 taxa in healthy controls). (**c**) CLR-normalized relative abundance values of water buffalo (*Bubalus bubalis*) DNA detected in stool samples compared across all timepoints. Box plots show median and interquartile ranges, while individual dots represent samples from each subject. Wilcoxon signed-rank test for 3-6 months vs 9 months and 10 months vs 14 months and Wilcoxon rank-sum test for all other timepoint comparisons were conducted. Significance results of statistical testing are marked. (**d**) CLR-normalized relative abundances of tea (*Camellia sinensis*) DNA amplified from stool samples compared across all timepoints. Box plots show median and interquartile ranges, while individual dots represent samples from each subject. *Wilcoxon signed-rank test* for 3-6 months vs 9 months and 10 months vs 14 months and *Wilcoxon rank-sum test* for all other timepoint comparisons were conducted. Significance results of statistical testing are marked. (**e**) Age-dependent increase in dietary diversity measured by traditional dietary diversity score (DDS, salmon) and plant FoodSeq Richness (pFR, cyan). Both metrics show significant positive correlation with age (p<0.0001; Linear mixed effects model with participant ID as random effect and PCR batch and gender as fixed effects). Shaded areas represent 95% confidence intervals.

To explore other dietary factors potentially influencing treatment outcomes, we analyzed broader FoodSeq patterns. Overall, 89 unique plant and 16 vertebrate taxa were identified (**Fig. 2b**). Wheat was detected across >50% of samples; banana was the most frequent fruit (31.6%); peas the most frequent legume (35%), and water buffalo the most common animal (35.8%) (**Fig. 2b**). Water buffalo further displayed consistently increasing CLR values from 3-6 months to 14 months (median= -0.68, IQR [0.36] to median= 3.26, IQR [5.79], *p* = 0.010; *Wilcoxon rank-sum test*) (**Fig. 2c**), aligning with the use of diluted water buffalo milk for complementary feeding reported in 24-hour recalls (data not shown). Notably, tea (*Camellia sinensis*), an age-inappropriate food per WHO infant feeding guidelines^25^, was present in >50% of samples and detected as early as 3–6 months (**Fig. 2d**). Tea CLR values rose significantly from 9 months to 10 months (median= 3.63, IQR [8.31] to median= 7.63, IQR [4.09], *p* = 0.006; *Wilcoxon rank-sum test*), then declined by 14 months (median= 5.76, IQR [3.59], *p* = 0.019; *Wilcoxon signed-rank test*) (**Fig. 2d**).

Next, to evaluate FoodSeq’s ability to capture expected dietary expansion with infant maturation, we measured both a traditionally calculated Dietary Diversity Score (DDS) and plant FoodSeq Richness (pFR), the number of unique identified plant taxa detected per sample^21^. Both metrics were positively correlated with age (β = 0.42, *p* < 0.0001 and β = 0.72, *p* < 0.0001 respectively) (**Fig. 2e**) but not with each other, suggesting capture of distinct aspects of dietary diversity. Consistent with prior findings from a global infant cohort, the primary axis of dietary variation (PC1) reflected total plant material from complementary feeding, as dietary loadings pointed almost exclusively in the same direction^26^ (**Fig. 2a**). In line with increased complementary feeding, PC1 scores increased with age, from 3-6 months to 10 months and 14 months, (*p* < 0.001; linear mixed effects model: gender + PCR batch as fixed effects, participant ID as random effect; healthy controls excluded) (**Fig. 2a**). Age also exerted a stronger influence on PC1 than health status (**Fig. S2a**).

Chickpea abundances at 10 months were not associated with DDS, pFR, or major dietary patterns (plant PCs) (**Fig. S2c**) and did not partition by wealth index, food insecurity, or family size (**Fig. S2b**), suggesting that RUSF compliance was not affected by local socio-economic factors or dietary practices. In the present context, in addition participant non-compliance, methodological factors such as a lag between RUSF consumption and stool collection might also contribute to the absence of chickpea DNA in some 10-month intervention samples (**Fig. 2b**).

Our findings highlight the potential of utilizing stool genomics to empirically capture geographic and cultural dietary patterns with minimal protocol modification through the universal language of DNA, augmenting CMAM interventions by tracking adherence and revealing contextual barriers to treatment. In our cohort, the most frequently detected taxa were staple crops in Pakistan and ingredients of common complementary foods: wheat in roti (flatbread) and daliya (porridge), rice and derivatives, and banana^17^, all verified from the 24-hr recalls as complementary foods reported in this cohort. Water buffalo, South Asia’s primary cattle species, is widely used for milk and meat^27^, underscoring the ability to taxonomically resolve regionally-specific sources of common foods. The consistent increase in detection of water buffalo DNA with age may further indicate weaning of infants from breast milk to cattle dairy and is supported by increased rates of dairy milk consumption reported across age in the 24-hr recalls. The widespread presence of tea DNA suggested a more extensive consumption during complementary feeding than previously reported (only one study documented use in >50% of infants), contrary to WHO guidelines^17^. However, neither tea nor other socioeconomic or dietary factors appeared to reduce RUSF intake, and chickpea-based RUSF was well-accepted for outpatient care, though measures to reduce tea provision may be warranted given the known impacts on appetite and iron absorption^17^.

Interestingly, though *Acha Mum* contains dried-milk, the pattern of milk-producing taxa abundances was not comparable to chickpea (**Fig. 2c**, **Fig. S3a**). This discrepancy may stem from DNA degradation during milk drying, low milk content in RUSF, or masking of RUSF-specific milk signal from milk in complementary feeding. These findings indicate a need to optimize detection methods for animal-derived RUSF/RUTF components. Chickpea DNA was detected in 22/30 samples from infants who received RUSF, yielding a sensitivity of 0.73 (specificity was not estimable due to absence of true negatives). The retrospective design, with spontaneous alignment of stool sampling to RUSF intake, likely contributed to non-detection, beyond inherent FoodSeq limitations. Future work should define the stool detection window post-RUSF/RUTF intake to guide sampling timing for improved sensitivity. Given FoodSeq’s potential to reflect portion size for certain plant taxa^21^, studies should quantify the relationship between RUSF/RUTF intake and residual DNA levels, and assess how formulations influence detectability across contexts. Though next-generation sequencing may be cost-prohibitive in low-resource settings, the use of ethanol-based preservation and room-temperature shipping^28^, along with targeted PCR assays and emerging portable PCR tools^29^, could lower costs and accelerate results. Lastly, stool collection faces barriers related to cultural sensitivities, logistics, and compliance; solutions may include community engagement and integration into existing healthcare workflows.

Overall, our study demonstrates the promise of FoodSeq as a novel tool for monitoring RUSF/RUTF compliance in the CMAM framework. Future trials could leverage FoodSeq to track adherence to other nutritional interventions or detect inappropriate feeding practices that compromise treatment. Standardizing ingredient detection, e.g., through marker food or synthetic DNA spike-ins, may enhance specificity and enable more precise quantification. As the method matures, FoodSeq insights could inform broader efforts to combat undernutrition, evaluate novel therapeutic foods, and shape child feeding policies.

## Methods

This study analyzed a subset of 90 participants from the Study of Environmental Enteropathy (SEEM; NCT03588013), a prospective longitudinal study conducted in rural Matiari, Pakistan (2016–2019), comprising malnourished (WHZ ≤ -2) and healthy (WHZ > 0, HAZ > -1.0) infants. Infants with malnutrition at 10 months received daily RUSF (*Acha Mum*: chickpea, oil, dried milk, sugar, micronutrients) for two months. Compliance was monitored through weekly home visits. Fecal samples were collected at 3–6, 9, 10, 14, and ≥12 months, flash-frozen without preservatives, shipped to Washington University in St. Louis, University, USA. Stool samples were pulverized and DNA extracted via bead beating with organic solvents. Samples were then shipped to Duke University, USA for FoodSeq analysis, a dual-marker amplicon sequencing approach amplifying plant (trnL-P6) and animal (12SV5) DNA, followed by Illumina library preparation and sequencing. Dietary DNA sequences were classified using curated reference databases and analyzed through DADA2 and phyloseq. Taxonomic read counts were center-log-ratio transformed, and taxa below threshold were filtered. Plant dietary compositionality was visualized using PCA, plant FoodSeq richness (pFR) was computed as previously described^21,28^, and dietary diversity was calculated across 19 food groups using 24-hour recalls. Statistical analyses included Wilcoxon tests and linear mixed-effect models adjusting for participant ID, sex, and amplification batch.

## Author contributions

A.A., N.T.I, S.A.A., and L.A.D conceptualized the study. A.A., N.T.I. S.I.S., S.A., F.U., K.A, M.J.B. performed data curation. A.A., S.I.S., S.A. conducted formal analysis. N.T.I., S.A.A., and L.A.D. provided funding for the study. A.A., T.K.M, O.O., N.I, N.T.I., S.A., F.U. performed experimental investigation. A.A., T.K.M., L.A.D., S.I.S. formulated methodology. A.A., S.J., and M.J.B. administered the project. L.A.D., N.T.I., A.A., M.J.B. and J.I.G provided resources. A.A. and T.K.M performed bioinformatic processing. L.A.D., N.T.I., S.A.A., and J.I.G supervised the project. A.A. created data visualizations. A.A. and S.I.S. wrote the original manuscript draft. A.A., L.A.D., N.T.I., S.A., K.A., M.J.B. reviewed and edited the final manuscript.

## Data availability

Deidentified data, data dictionaries, code notebooks, and clinical metadata for reproducing manuscript results from processed *trnL-P6* and *12SV5* data can be provided upon request and will be shared in compliance with relevant study agreements, ethical standards, and regulations. With regards to *12SV5* data, a synthetic DNA sequence was generated to match the length and read count distribution of the original human mitochondrial sequences. This sequence does not correspond to any real individual and is provided solely to enable replication of the data analysis pipeline without compromising participant privacy.

## Acknowledgements

*Matiari* data management team and field staff for study support; families and their children for study participation; Marty Meier for technical assistance in purifying DNA from fecal samples; Michelle Kirtley and Anna Bauer for editorial guidance; Tonya Snipes, Lisa Alston-Latta, Joey Biondo, and Christopher Sutton for lab space maintenance and cleaning.

## Extended Methods

This study included a subset of 90 participants from the Study of Environmental Enteropathy (SEEM; NCT03588013; 365 malnourished with WHZ ≤ -2, 51 healthy with WHZ > 0, HAZ > −1.0), a prospective longitudinal study conducted in rural *Matiari*, Pakistan (March 2016-March 2019) ^18^ (**Fig. 1a**; [Created in BioRender. Aqeel, A. (2022) https://BioRender.com/f56s058Wti]). SEEM followed infants from birth to 24 months through a multi-national collaboration between Aga Khan University, Washington University in St. Louis, University of Virginia, and Cincinnati Children’s Hospital Medical Center, with funding from the Bill and Melinda Gates Foundation ^18^. Our subset comprised three groups: 30 malnourished infants with paired samples at 3-6 and 9 months, 30 malnourished infants with paired samples at 10-14 months, and 30 healthy controls sampled after 12 months of age. Caregivers of malnourished infants received complementary feeding education at enrollment (3-6 months), with anthropometric measurements recorded monthly ^18^. Infants maintaining WHZ < -2 at 9 months received a two-month intervention of daily *Acha Mum*, a locally-produced ready-to-use supplementary food (RUSF; ingredients: chickpea, edible oil, dried milk, sugar, vitamins [A, B1, B2, B3, B5, B6, B9, B12, C, D, E, K, H], zinc, folate, iodine, calcium etc. emulsifier) ^18^. Study staff monitored compliance through weekly home visits, with additional visits as needed ^18^. A previous Phase 1 study demonstrated 90.5% intervention adherence ^18^. The RUSF intervention was administered for 57.8 ± 6.3 days beginning at 9.89 ± 0.99 months ^30^. Non-responders (WHZ < -2 post-intervention) without evidence of celiac disease or other identifiable growth failure etiologies underwent esophagogastroduodenoscopy (EGD) for investigation of underlying pathophysiology ^18^. Compliance was monitored through weekly home visits by study staff ^18^.

Fecal samples were collected by CHWs (approximately 1 g per participant per timepoint) within 10 minutes of production and transferred to liquid nitrogen pre-charged dry shippers without additives or preservatives (Taylor Wharton, CX-100) for transfer to and storage at -80°C at Aga Khan University (AKU) research facilities prior to shipping on dry ice to Washington University in St. Louis (WUSTL), USA. Sample collection occurred at 3-6, 9, 10, and 14 months for malnourished infants and at ≥12 months for healthy controls. At WUSTL, frozen fecal samples underwent pulverization in liquid nitrogen using a sterile mortar and pestle. A 50 mg aliquot of pulverized material was transferred to a 2 mL screw-cap vial containing. A solution consisting of 500 µL of phenol: chloroform: isoamyl alcohol (25:24:1), 210 µL of 20% SDS and 500 µL of buffer (200 mM NaCl, 200 mM Trizma base, 20 mM EDTA) containing 500 µL of 0.1 mm diameter zirconia/silica beads was added to each sample, followed by bead beating (Mini-Beadbeater-8; Biospec) to extract DNA. DNA was purified (Qiaquick columns, Qiagen), eluted in 70 µL of Tris-EDTA (TE) buffer, and quantified (Quant-iT dsDNA broad range kit; Invitrogen).

Residual dietary plant DNA and animal DNA in stool samples was assessed using FoodSeq, as previously described ^21,24^. Briefly, FoodSeq libraries were generated using a two-step PCR protocol. Primary amplification was conducted using locus-specific primers (trnL *g-h* for plants and 12SV5F/12SV5R76 for animals) with Illumina overhang adapter sequences, a human blocking primer (DeBarba14 HomoB77 for 12SV5), and SYBR Green for qPCR. PCR reactions were performed in a total volume of 10 µl using either AccuStart II PCR SuperMix (12SV5) ^24^ or previously established conditions (trnL) ^21^. Cycling conditions included an initial denaturation at 94°C for 3 minutes, followed by 35 cycles of denaturation, annealing, and extension. Each PCR batch included positive and negative controls, and batches were repeated if controls failed ^24^. Secondary PCR was conducted in a 50 µl reaction volume using KAPA HiFi polymerase to add Illumina adapters and 8 bp dual indices for sample multiplexing ^24^. The final libraries were cleaned, pooled separately for trnL and 12SV5, and sequenced on independent Illumina runs ^24^.

Curated reference databases were constructed by compiling a list of edible plant and animal taxa from global food surveys and reference sources ^24^. Sequences containing trnL or 12SV5 regions were retrieved from GenBank and RefSeq, filtered for primer binding sites with a ≤20% mismatch tolerance, and trimmed to the target amplicon regions ^24^. Identical sequences from different accessions were de-duplicated, while intra-taxon variability and conserved sequences across taxa were retained ^24^. Demultiplexed reads were processed using BBDuk (adapter trimming), Cutadapt (primer filtering and additional trimming), and DADA2 (denoising, merging, and amplicon sequence variant (ASV) inference) ^24^. Reads were quality-filtered using an expected error threshold of 2 and truncated at the first base with a quality score ≤2 ^24^. For trnL, ASVs were assigned using DADA2’s assignSpecies function, with exact sequence matching to the custom trnL reference database ^24^. In cases of ambiguous matches, taxa were assigned to the last common ancestor (e.g., a sequence matching both wheat and rye was classified at the Poaceae family level) ^24^. For 12SV5, ASVs were assigned using DADA2’s assignTaxonomy and glommed at the lowest common taxonomic assignment, due to reduced performance of exact sequence matching caused by PCR polymerase-induced mismatches^24^. Suspected contaminants were identified using decontam and removed based on DNA quantitation data ^24^. ASV count tables, taxonomic assignments, and metadata were structured using phyloseq (version 1.38.0) ^24^.

For statistical analyses, taxonomic read counts were converted to relative abundances and normalized through center-log-ratio transformation to preserve overall data structure followed by removal of taxa with fewer than 5 sequence reads, removal of taxa with no assignment at the superkingdom level in trnL and removal of human ASVs from the 12SV5 data. Plant dietary compositionality was analyzed using Principal Component Analysis (PCA, *’prcomp’* function, parameters: centered, not scaled). Plant FoodSeq richness (pFR) was calculated as the number of unique taxa with at least one read count in each sample as previously described ^21^. The Dietary Diversity Score (DDS) was derived from 24-hr dietary recall data ^31^. The data collection process, as well as the complete methodology for standardization and homogenization has been previously described ^31^. The reported food items were categorized into 19 predefined food groups following a standardized food classification system ^31^. Food groups considered included: cereals and their products; pulses, legumes, and their products; nuts, seeds, and their products; leafy vegetables and their products; vegetables and their products; starchy roots and tubers; flesh meat and their products; fish, seafood, and their products; organ meats; fruits and their products; beverages; milk and their products; sweets and their products; spices, condiments, and seasoning foods; fats and oils; non-food items; supplements; and eggs and their products. Although standard DDS calculations incorporate up to ten food groups, for this study we classified food in greater detail for improved comparison with genomic dietary data resolution. Each participant’s dietary intake was assessed to determine whether at least one item from each food group was consumed within the recall period. A binary scoring system was applied, where the presence of a food group was assigned a score of ‘1’, while its absence was assigned a score of ‘0’. The DDS was calculated as the sum of all consumed food groups:

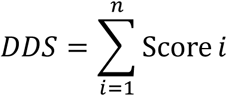

where Score represents the presence (1) or absence (0) of the *i*th food group, and n is the total number of food groups considered (19 in this study). In terms of comparative testing, *Wilcoxon rank-sum test* was used for comparisons between unpaired samples and *wilcoxon signed-rank test* for paired samples. Relationships spanning all timepoints were assessed using linear mixed effects models with participant ID as a random effect and gender and PCR batch as additional fixed effects.

## Supplemental Figures

**Fig. S1.**
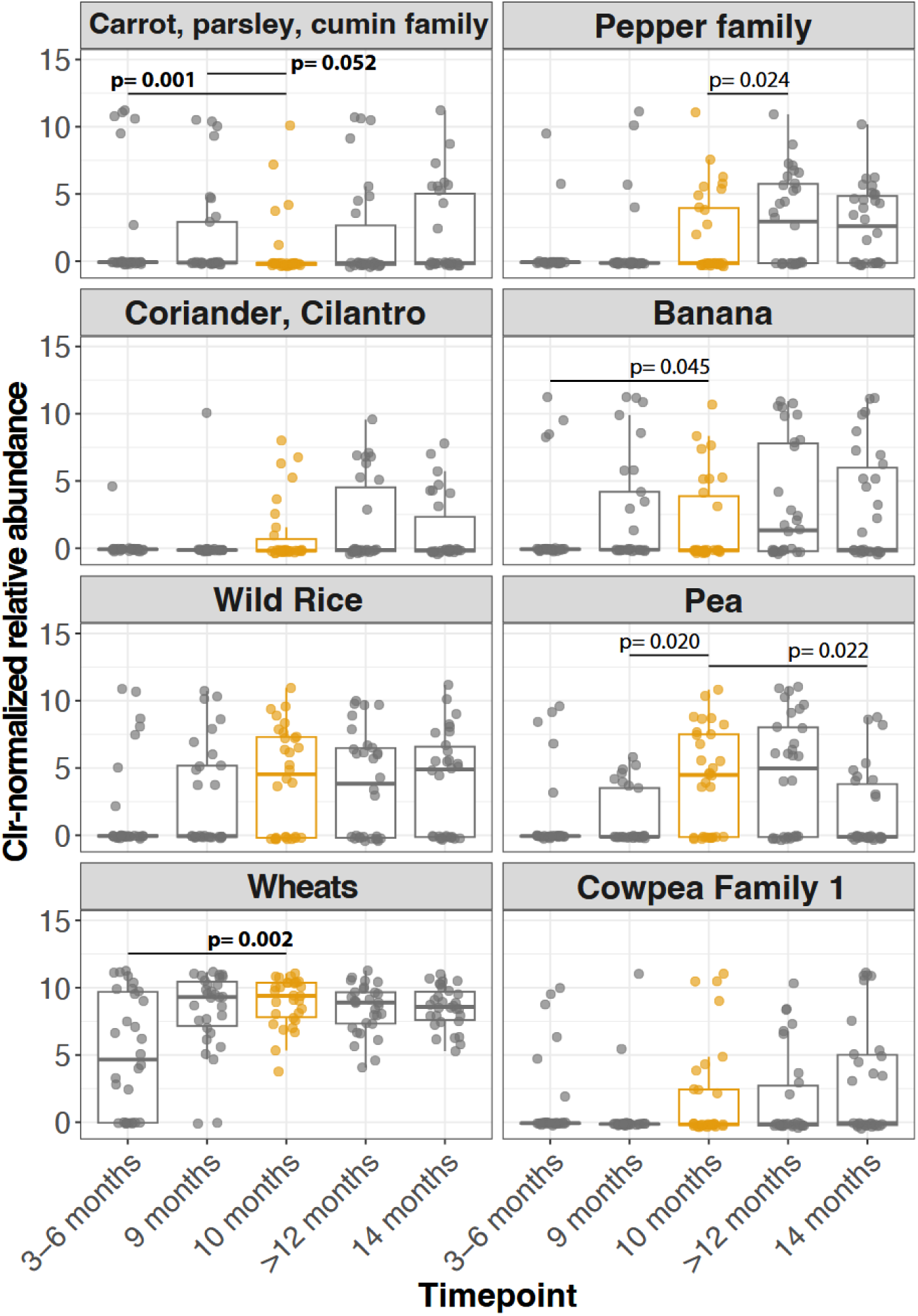
Major dietary taxa DNA detection patterns do not align with RUSF administration. CLR-normalized DNA relative abundances of top 10 taxa driving dietary composition as identified. By PCA biplot compared across all timepoints. The 10-month timepoint when RUSF was administered is highlighted in orange. Box plots show median and interquartile ranges, while individual dots represent samples from each subject. *Wilcoxon signed-rank test* for 3-6 months vs 9 months and 10 months vs 14 months and *Wilcoxon rank-sum test* for all other timepoint comparisons were conducted. Significance results of statistical testing are marked. Significance values in bold remain significant after Benjamini-Hochberg multiple testing correction (including Chickpea and Tea timepoint comparison tests).

**Table S1.**
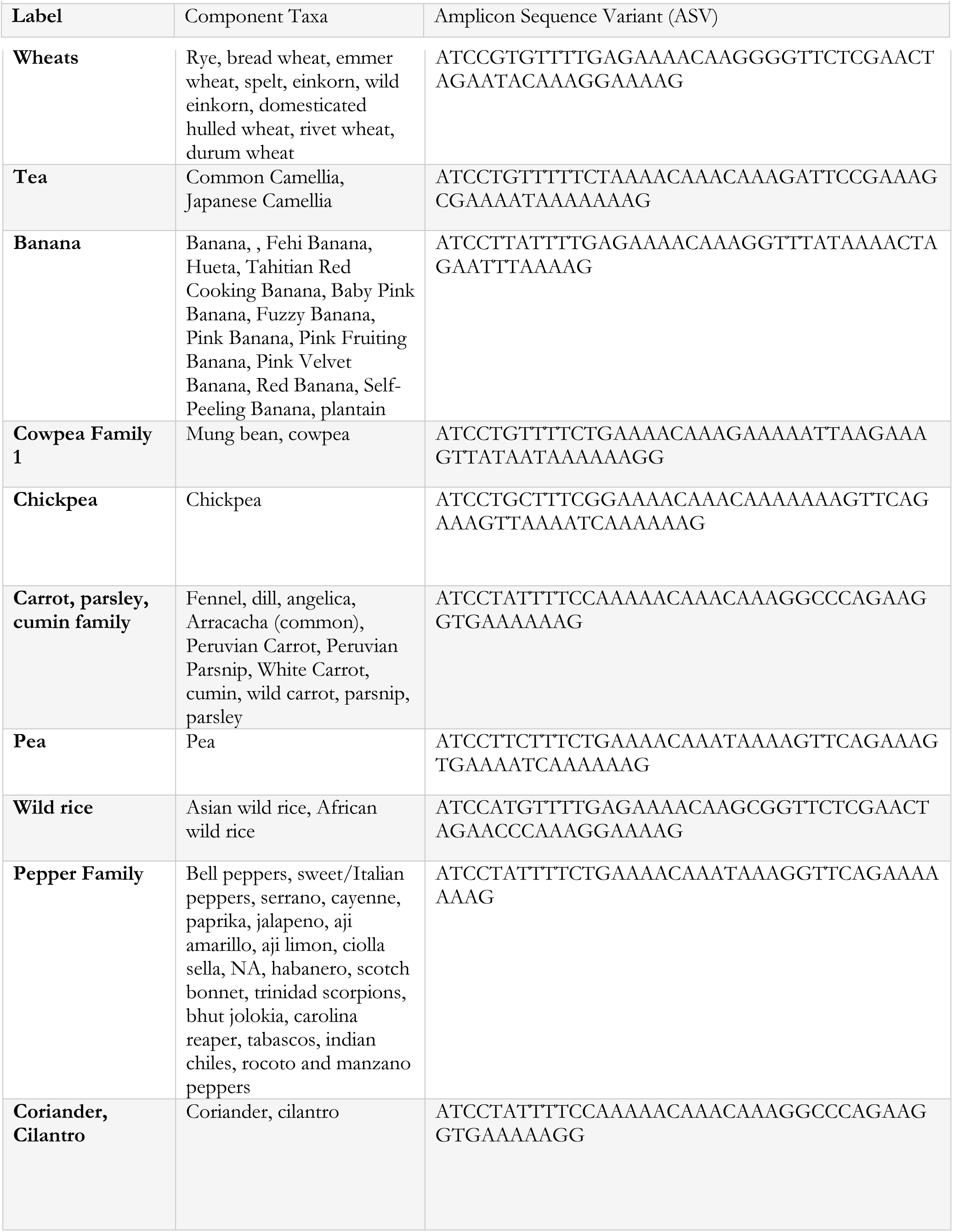
Top taxa identified as drivers of PC1 and PC2 in dietary principal components analysis.

**Fig. S2.**
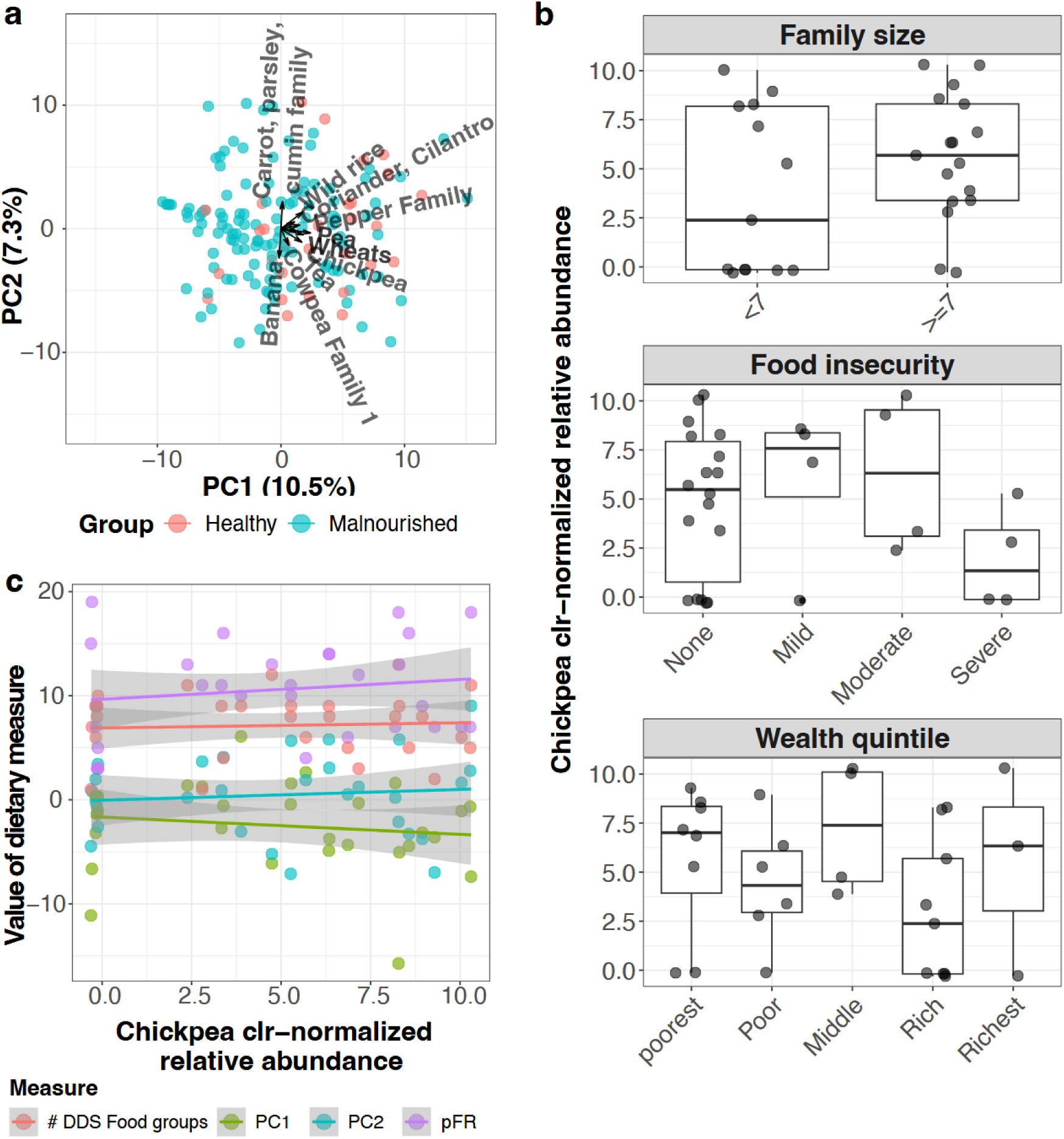
Health status does not impact dietary composition and chickpea DNA detection is not impacted by socioeconomic or other dietary factors. (**a**) Principal Component Analysis (PCA) of plant dietary composition with biplot indicating relative contributions of top 10 dietary components colored by health status (healthy controls vs malnourished samples) and clusters denoted by ellipses. Health status compared along PC1 and PC2 values using linear mixed effect models with age, gender, and PCR batch as fixed effects and sample ID as random effects. No significant impact detected when accounting for age. (**b**) Chickpea CLR-normalized relative abundances at the 10-month timepoint compared across family size, food insecurity status, wealth quintile of surveyed households using *Wilcoxon rank-sum test*. Box plots show median and interquartile ranges, while individual dots represent samples from each subject. No significant difference detected. (**c**) Chickpea CLR-normalized relative abundances compared at the 10-month timepoint against dietary factors such as traditional and genomic dietary diversity as well as the first two principal components from a plant dietary composition PCA excluding chickpea (each PC approximating a broader dietary pattern). Linear mixed effects model with participant ID as random effect and PCR batch, and gender as fixed effects. Shaded areas represent 95% confidence intervals. No significant relationship detected.

**Fig. S3.**
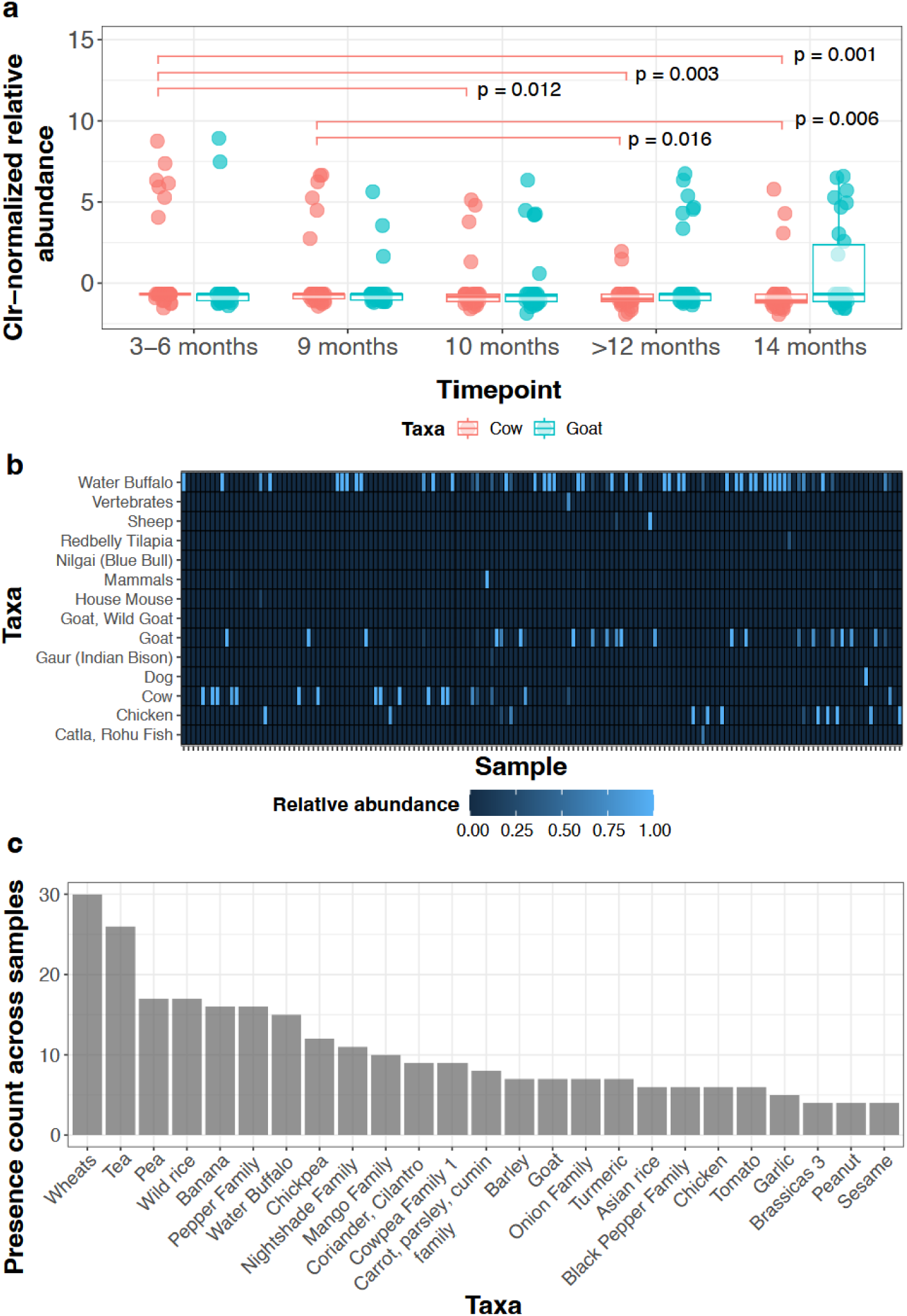
Dietary DNA Composition in Stool Samples: Heatmap of Vertebrate Taxa Abundances and Presence Counts of Common Food Taxa in Healthy Controls. (**a**) Clr-normalized relative abundance values of cow and goat DNA detected in stool samples compared across all timepoints. Box plots show median and interquartile ranges, while individual dots represent samples from each subject. Wilcoxon signed-rank test for 3-6 months vs 9 months and 10 months vs 14 months and Wilcoxon rank-sum test for all other timepoint comparisons were conducted. Significance results of statistical testing are marked. (**b**) Heatmap of relative abundances of vertebrate taxa DNA post-filtration of human DNA and taxa with less than 5 total reads. Low-frequency detection of likely environmental contaminants (e.g. dog, Indian mouse) may reflect poor sanitation or food contamination. (**c**) Bar plot showing the presence counts of top 25 food (plant and vertebrate combined) taxa detected in participant stool samples in healthy controls (>12 months).

